# Mass drug administration with azithromycin for trachoma elimination and the population structure of Streptococcus pneumoniae in the nasopharynx

**DOI:** 10.1101/2020.04.01.20047266

**Authors:** Rebecca A. Gladstone, Ebrima Bojang, John Hart, Emma M Harding-Esch, David Mabey, Ansumana Sillah, Robin L. Bailey, Sarah E. Burr, Anna Roca, Stephen D. Bentley, Martin J. Holland

**Author notes:** Corresponding author: Rebecca. A. Gladstone, Infection genomics, Wellcome Sanger Institute, Hinxton, England, UK,. Alternative corresponding author: Martin J Holland, Clinical Research Department, London School of Hygiene and Tropical Medicine, Keppel Street, London, England, UK.

## Abstract

**Background:** Mass drug administration (MDA) with azithromycin for trachoma elimination reduces nasopharyngeal carriage of *Streptococcus pneumoniae* in the short term. We evaluated S. *pneumoniae* carried in the nasopharynx before and after a round of azithromycin MDA to determine whether MDA was associated with changes in pneumococcal population structure.

**Methods:** We analysed 514 pneumococcal isolates cultured from nasopharyngeal samples collected in Gambian villages that received MDA for trachoma elimination. The samples were collected during three cross-sectional surveys conducted before the third round of MDA (CSS-1) and at one (CSS-2) and six (CSS-3) months after MDA. Whole genome sequencing was conducted on randomly selected isolates. Bayesian Analysis of Population Structure (BAPS) was used to cluster related isolates by capturing variation in the core genome. Serotype and multi-locus sequence type were inferred from the genotype. The Antimicrobial Resistance Identification by Assembly (ARIBA) tool was used to identify macrolide resistance genes.

**Results:** Twenty-seven BAPS clusters were assigned. These consisted of 81 sequence types (STs), 15 of which were novel additions to pubMLST. Two BAPS clusters, BAPS20 (p-value<=0.016) and BAPS22 (p-value<=0.032) showed an increase in frequency at CSS-3 not associated with antimicrobial resistance. Macrolide resistance within BASP17 increased after treatment (p<0.05) and was carried on a mobile transposable element that also conferred resistance to tetracycline.

**Conclusions:** Limited changes in pneumococcal population structure were observed after the third round of MDA suggesting treatment had little effect on the circulating lineages. An increase in macrolide resistance within one BAPS highlights the need for antimicrobial resistance surveillance in treated villages.

## INTRODUCTION

Trachoma, caused by ocular infection with *Chlamydia trachomatis, remains* the leading infectious cause of blindness worldwide. One facet of the World Health Organization (WHO)-recommended strategy for trachoma elimination involves mass drug administration (MDA) with the broad-spectrum antibiotic azithromycin^1,2^. This serves to decrease the reservoir of infection, thereby reducing transmission.

Azithromycin MDA, given in the context of trachoma control, has also been shown to have important collateral benefits including reduced child mortality ^3-5^. This was first documented in Ethiopian villages where the mortality rate of children aged 1 to 9 years and residing in treated villages was lower than that in untreated villages ^3^. More recently, a trial conducted in Niger, Tanzania and Malawi found biannual MDA with azithromycin significantly reduced all cause under-5 mortality in comparison to a placebo ^5^. While the mechanism underlying this reduction is not yet clear it may be due, in part, to a reduction in carriage of *Streptococcus pneumoniae*. Indeed, a single round of azithromycin MDA, given as a part of trachoma elimination programmes, has been consistently shown to significantly reduce prevalence of S. *pneumoniae* carriage at the community level ^6-9^.

The effect of MDA on S. *pneumoniae* carriage is short-lived with rates returning to original levels within a few months of treatment ^10^. However, it is unclear from the studies conducted to date whether the S. *pneumoniae* strains that are circulating within the population before treatment are the same as those that are found when carriage rates return to their previous levels after treatment, or whether azithromycin MDA selects for previously suppressed strains or enables clonal expansion. One study, conducted in Ethiopia and following intensive MDA given at three-monthly intervals over the course of one year, documented decreased serotype diversity and was associated with a significant rise in macrolide resistance following treatment. The authors of the study concluded that clonal expansion of resistant isolates caused a decrease in the diversity of the population ^11^.

Whole genome sequencing data allows high-resolution definitions of clones to be made. Bayesian Analysis of Population Structure (BAPS) has been used to define the population structure of pneumococcal collections in a number of studies assessing the impact of medical inventions ^12-14^. Rather than relying on serotype, which can be expressed in multiple genetic backgrounds and/or variation in the seven multi-locus-sequence-typing (MLST) genes, BAPS uses variation across the genome to cluster a population into closely related groups of isolates. BAPS clusters typically aggregate closely related MLST sequence types (STs) and can be used to assess any restructuring of the pneumococcal population, with increased the power to detect small changes in prevalence across multiple sub-clones.

In The Gambia, we have previously shown that azithromycin MDA resulted in a significant yet transient decrease in S. *pneumoniae* carriage with prevalence falling from 43% before a third and final round of MDA to just 19% one month following MDA [9]. Within six months of MDA, prevalence of carriage returned to 45% but was not associated with a significant rise in the carriage of azithromycin resistant strains. The aim of the present study was to determine if there were changes in S. *pneumoniae* population structure following the third round of MDA. To address this aim we randomly selected isolates taken from the carriage study and conducted whole genome sequencing which were then examined using BAPS to infer population structure.

## METHODS

### Study design

The Partnership for the Rapid Elimination of Trachoma (PRET) study (ClinicalTrials.gov NCT00792922) was a cluster randomized controlled trial, the design of which has been described elsewhere ^15-17^. Briefly, the study compared the effectiveness of three annual rounds versus one round of azithromycin MDA in reducing the prevalence of active trachoma and ocular C. trachomatis infection. All residents of a community were eligible for treatment, given as a single oral dose of 20 mg azithromycin per kg to a maximum of 1 g. Height was used as a proxy for weight. A pneumococcal carriage study was nested within PRET ^9^ and was carried out in eight villages that were part of the larger trial. This included two villages that had been randomized (by the underlying PRET trial) to three annual rounds of MDA (3×treatment arm) and six villages that received a single treatment round (1×treatment arm). All villages had also participated in a cluster (by village) randomized trial of a 7-valent pneumococcal vaccine (PCV-7) ^16^. Villages selected for the study presented here were part of the trial’s control arm in which children under 5 years of age and those born during the trial period had received 1-3 vaccine doses. The trial took place between 2006 and 2008, before PCV-7 introduction in The Gambia. PCV-7 modified distribution of pneumococcal serotypes in carriage with a strong decrease of serotypes included in the vaccine in all age groups.

During the course of the pneumococcal carriage study, three cross-sectional surveys (CSS) were conducted in the 3×treatment arm of PRET: CSS-1, one month prior to the third annual round of MDA; CSS-2, one month following the third round of MDA; and CSS-3, 6 months following the third round (Figure 1). Census data were gathered in the week prior to the onset of the study. At CSS-1, all censused children <15 years of age, present at the time of sampling were invited to participate. For individuals ≥15 years, 150 were randomly selected for participation. At each of CSS-2 and CSS-3, participation was restricted to those who had received azithromycin during the third round of MDA in July 2010; all censused and treated children <15 years of age were invited to participate while 150 treated individuals ≥15 years were randomly selected for participation.

**Figure 1.**
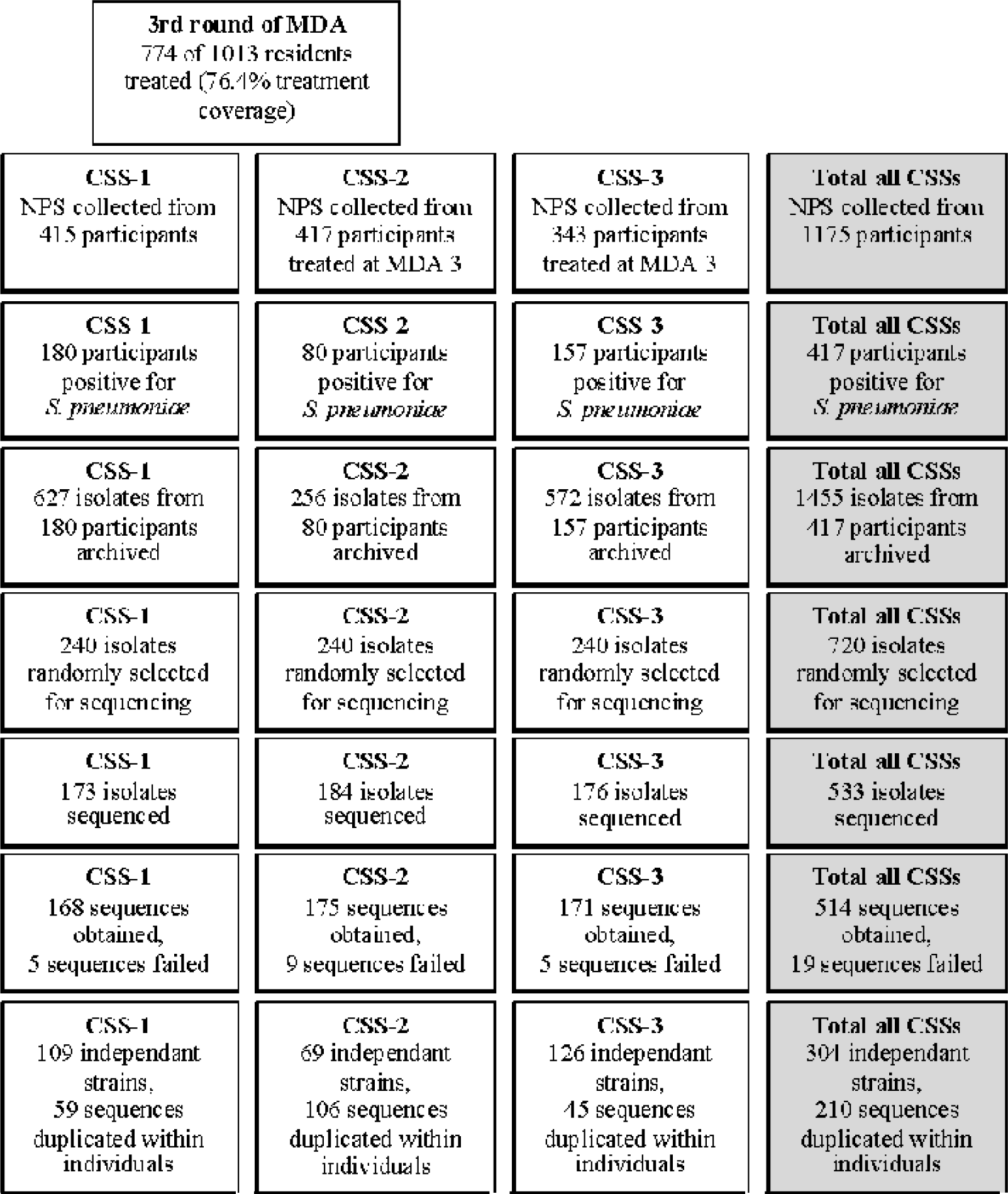
Study flow.

In the current study, 720 S. *pneumoniae* isolates, out of 1455 archived during these CSSs, were randomly selected for whole genome sequencing. Randomization was restricted such that CCS1 to 3 would be equally represented by children less than 5 years of age, by children between 5 and 10 years of age and by participants over 10 years of age. The 240 isolates from each CSS were randomly selected by assigning all isolates from that CSS in the archive a random number. These random numbers were then ordered from largest to smallest and the top 240 were selected.

### Isolate selection, culture conditions and DNA extraction

During the original pneumococci carriage study, up to 4 S. *pneumoniae* isolates representing varying colony morphologies were cultured from each positive nasopharyngeal swab using conventional microbiological techniques as detailed in Burr et al.^9^. Isolates randomly selected for whole genome sequencing were revived from glycerol stocks maintained at -70⍰C. All isolates were regrown on blood agar plates containing gentamicin and incubated overnight at 37⍰C with 5% CO_2_. The following day, a single well-isolated colony was inoculated into 20 ml Todd-Hewitt broth containing 0.5% yeast extract and re-incubated overnight under the same conditions. The overnight broth cultures were pelleted by centrifugation and DNA was extracted using the Wizard Genomic DNA Purification Kit according to the manufacturer’s protocol (Promega Corporation, Madison, WI, USA). DNA quantification was performed on all extracts using PicoGreen staining (Invitrogen, Paisley, UK) and those samples for which a minimum of 500 ng DNA had been obtained were forwarded for sequencing.

### Sequencing and sequence analysis

DNA was sequenced on Illumina Hiseq platforms using pair-end reads lengths of 100 base pairs. 595 isolates were sequenced and of those 514 were confirmed as pneumococcal streptococci, had sufficient coverage and were not heavily contaminated with other species. Core genes were defined using Roary ^18^. The core alignment was clustered using hierBAPS^19^ to determine BAPS clusters, with a second level of hierarchical clustering to determine BAPS sub-clusters. The core alignment was reduced to Single Nucleotide Polymorphism (SNP)-sites^20^ and was used to produce a phylogenetic tree using RAxML ^21^, midpoint rooted, in the absence of classical non-typeable isolates to serve as a natural outgroup. SNPs were reconstructed on the tree using the acctran parsimony method. Serotypes were inferred from the genomic data using a mapping-based technique ^22^. The Antimicrobial Resistance Identification by Assembly (ARIBA) tool ^23^ and a pneumococcal typing tool ^24^ were used to screen all isolates identifying known genes and mutations that confer resistance, including ermB and mef that explain almost all macrolide resistance in pneumococci ^24^.

### Statistical analysis

Only one isolate of the same sequence type (ST) derived from the same individual at the same timepoint was included in the statistical analysis 304/514. The diversity of serotypes and STs was assessed using Simpson’s Diversity index (Dominance Index Approximation), which represents the probability that two randomly selected strains are the same, where 1 means they are different and 0 indicates they are the same.

The distribution of a particular serotype was calculated as the number of strains belonging to that serotype at a given CSS divided by the total number of analysed strains at that CSS. Confidence intervals on the observations sampled were calculated at 95% using the R Hmisc binconf “Confidence Intervals for Binomial Probabilities” with the exact method. Statistically significant changes in the proportion of a particular BAPS, serotype or ST between two time-points was performed if at least five isolates represented the serotype/genotype being tested. These changes were determined using Fisher’s exact test to produce a two-tailed p-value in R adjusted for multiple testing using p.adust with “BH” method.

### Sample size

The study’s primary objective was to determine whether particular BAPS clusters increased in frequency following MDA. The number of independent isolates for which quality sequence data was obtained (304) provided 80% power with 95% confidence to detect a five-fold increase of one of these clusters between CSS-1 and CSS-2 and four-fold increase between CCS-1 and CSS-3.

Metadata including ENA accession numbers and phylogeny are deposited in Microreact https://microreact.org/project/MDAandpneumo.

### Ethical review

The study adhered to the tenants of the Declaration of Helsinki and was approved by The Gambia Government/Medical Research Council Unit, The Gambia Joint Ethics Committee. Written, informed consent was obtained from all participants. In the case of minors, informed consent was obtained from the parent or guardian.

## RESULTS

The study flow is shown in Figure 1. During the initial carriage study, 1,455 isolates were generated from participants in the 3×treatment arm. Of these, 720 were randomly selected for whole genome sequencing, 240 from each CSS, such that each CCS would be equally represented by children less than 5 years of age, by children between 5 and 10 years of age and by participants over 10 years of age. 533 of these 720 isolates were successfully revived from glycerol stocks and had subsequent overnight cultures that yielded at least 500 ng, which was required for DNA sequencing. Quality sequence data was obtained for 514 of these isolates. Of those sequences that failed quality control, 1 had very low sequence yield, and 18 showed evidence of contamination. Excluding multiple instances of a ST from the same participant at the same time point (210 instances), 304 independent isolates were available to assess temporal distribution and diversity (Figure 1).

### Bayesian Analysis of Population Structure

BAPS revealed 27 BAPS clusters (Figure 2). A comparison of the proportion of isolates corresponding to a particular cluster at CSS-1 and CSS-2 identified no BAPS clusters as having changed significantly in proportion in the month following MDA (Table 1). Significant changes in BAPS clusters between CSS-1 and CSS-3 were observed with BAPS20 (p=0.016) and BAPS22 (p=0.032). Neither of these BAPS was observed at CSS-1 or CSS-2 but both were found at CSS-3, six months following MDA [BAPS20, 8.73% sequenced isolates at CSS-3 (95% CI 4.44-15.08); BAPS22, 7.14% sequenced isolates at CSS-3 (95% CI 3.32-13.13)] and were comprised exclusively of serotype 16F (ST8949/13858) and 45 (ST2831), respectively.

**Table 1.**
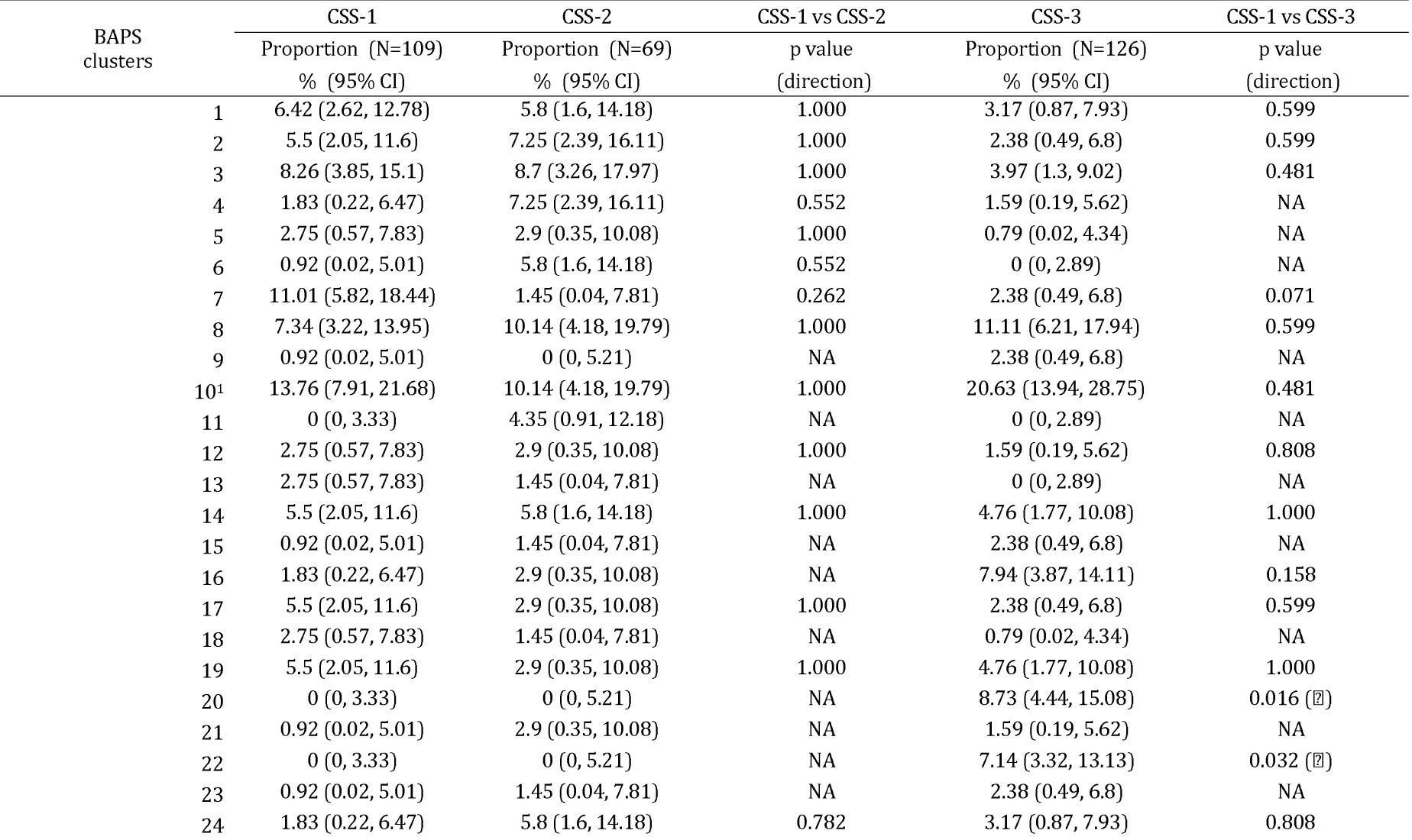
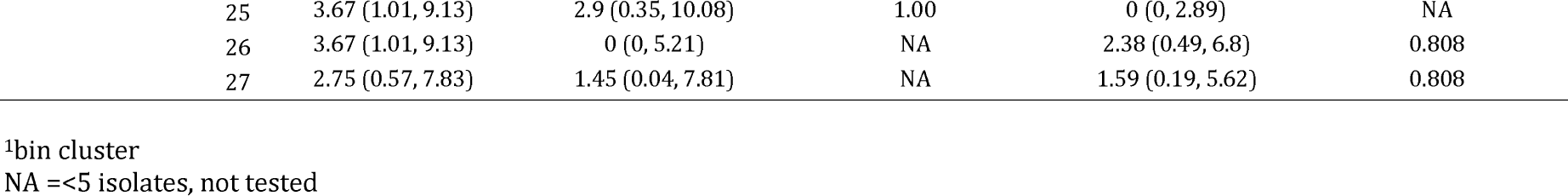
Proportion of isolates belonging to BAPS clusters at each CSS.

| BAPS<br>clusters |  | CSS-1 | CSS-2 | CSS-1 vs CSS-2 | CSS-3 | CSS-1 vs CSS-3 |
| --- | --- | --- | --- | --- | --- | --- |
|  |  | Proportion (N=109) | Proportion (N=69) | p value | Proportion (N=126) | p value |
|  |  | % (95% CI) | % (95% CI) | (direction) | % (95% CI) | (direction) |
|  | 1 | 6.42 (2.62, 12.78) | 5.8 (1.6, 14.18) | 1.000 | 3.17 (0.87, 7.93) | 0.599 |
|  | 2 | 5.5 (2.05, 11.6) | 7.25 (2.39, 16.11) | 1.000 | 2.38 (0.49, 6.8) | 0.599 |
|  | 3 | 8.26 (3.85, 15.1) | 8.7 (3.26, 17.97) | 1.000 | 3.97 (1.3, 9.02) | 0.481 |
|  | 4 | 1.83 (0.22, 6.47) | 7.25 (2.39, 16.11) | 0.552 | 1.59 (0.19, 5.62) | NA |
|  | 5 | 2.75 (0.57, 7.83) | 2.9 (0.35, 10.08) | 1.000 | 0.79 (0.02, 4.34) | NA |
|  | 6 | 0.92 (0.02, 5.01) | 5.8 (1.6, 14.18) | 0.552 | 0 (0, 2.89) | NA |
|  | 7 | 11.01 (5.82, 18.44) | 1.45 (0.04, 7.81) | 0.262 | 2.38 (0.49, 6.8) | 0.071 |
|  | 8 | 7.34 (3.22, 13.95) | 10.14 (4.18, 19.79) | 1.000 | 11.11 (6.21, 17.94) | 0.599 |
|  | 9 | 0.92 (0.02, 5.01) | 0 (0, 5.21) | NA | 2.38 (0.49, 6.8) | NA |
|  | 10<sup>1</sup> | 13.76 (7.91, 21.68) | 10.14 (4.18, 19.79) | 1.000 | 20.63 (13.94, 28.75) | 0.481 |
|  | 11 | 0 (0, 3.33) | 4.35 (0.91, 12.18) | NA | 0 (0, 2.89) | NA |
|  | 12 | 2.75 (0.57, 7.83) | 2.9 (0.35, 10.08) | 1.000 | 1.59 (0.19, 5.62) | 0.808 |
|  | 13 | 2.75 (0.57, 7.83) | 1.45 (0.04, 7.81) | NA | 0 (0, 2.89) | NA |
|  | 14 | 5.5 (2.05, 11.6) | 5.8 (1.6, 14.18) | 1.000 | 4.76 (1.77, 10.08) | 1.000 |
|  | 15 | 0.92 (0.02, 5.01) | 1.45 (0.04, 7.81) | NA | 2.38 (0.49, 6.8) | NA |
|  | 16 | 1.83 (0.22, 6.47) | 2.9 (0.35, 10.08) | NA | 7.94 (3.87, 14.11) | 0.158 |
|  | 17 | 5.5 (2.05, 11.6) | 2.9 (0.35, 10.08) | 1.000 | 2.38 (0.49, 6.8) | 0.599 |
|  | 18 | 2.75 (0.57, 7.83) | 1.45 (0.04, 7.81) | NA | 0.79 (0.02, 4.34) | NA |
|  | 19 | 5.5 (2.05, 11.6) | 2.9 (0.35, 10.08) | 1.000 | 4.76 (1.77, 10.08) | 1.000 |
|  | 20 | 0 (0, 3.33) | 0 (0, 5.21) | NA | 8.73 (4.44, 15.08) | 0.016 (↗) |
|  | 21 | 0.92 (0.02, 5.01) | 2.9 (0.35, 10.08) | NA | 1.59 (0.19, 5.62) | NA |
|  | 22 | 0 (0, 3.33) | 0 (0, 5.21) | NA | 7.14 (3.32, 13.13) | 0.032 (↗) |
|  | 23 | 0.92 (0.02, 5.01) | 1.45 (0.04, 7.81) | NA | 2.38 (0.49, 6.8) | NA |
|  | 24 | 1.83 (0.22, 6.47) | 5.8 (1.6, 14.18) | 0.782 | 3.17 (0.87, 7.93) | 0.808 |

|  |  |  |  |  |  |
| --- | --- | --- | --- | --- | --- |
| 25 | 3.67 (1.01, 9.13) | 2.9 (0.35, 10.08) | 1.00 | 0 (0, 2.89) | NA |
| 26 | 3.67 (1.01, 9.13) | 0 (0, 5.21) | NA | 2.38 (0.49, 6.8) | 0.808 |
| 27 | 2.75 (0.57, 7.83) | 1.45 (0.04, 7.81) | NA | 1.59 (0.19, 5.62) | 0.808 |

<sup>1</sup>bin cluster
NA =<5 isolates, not tested

**Figure 2.**
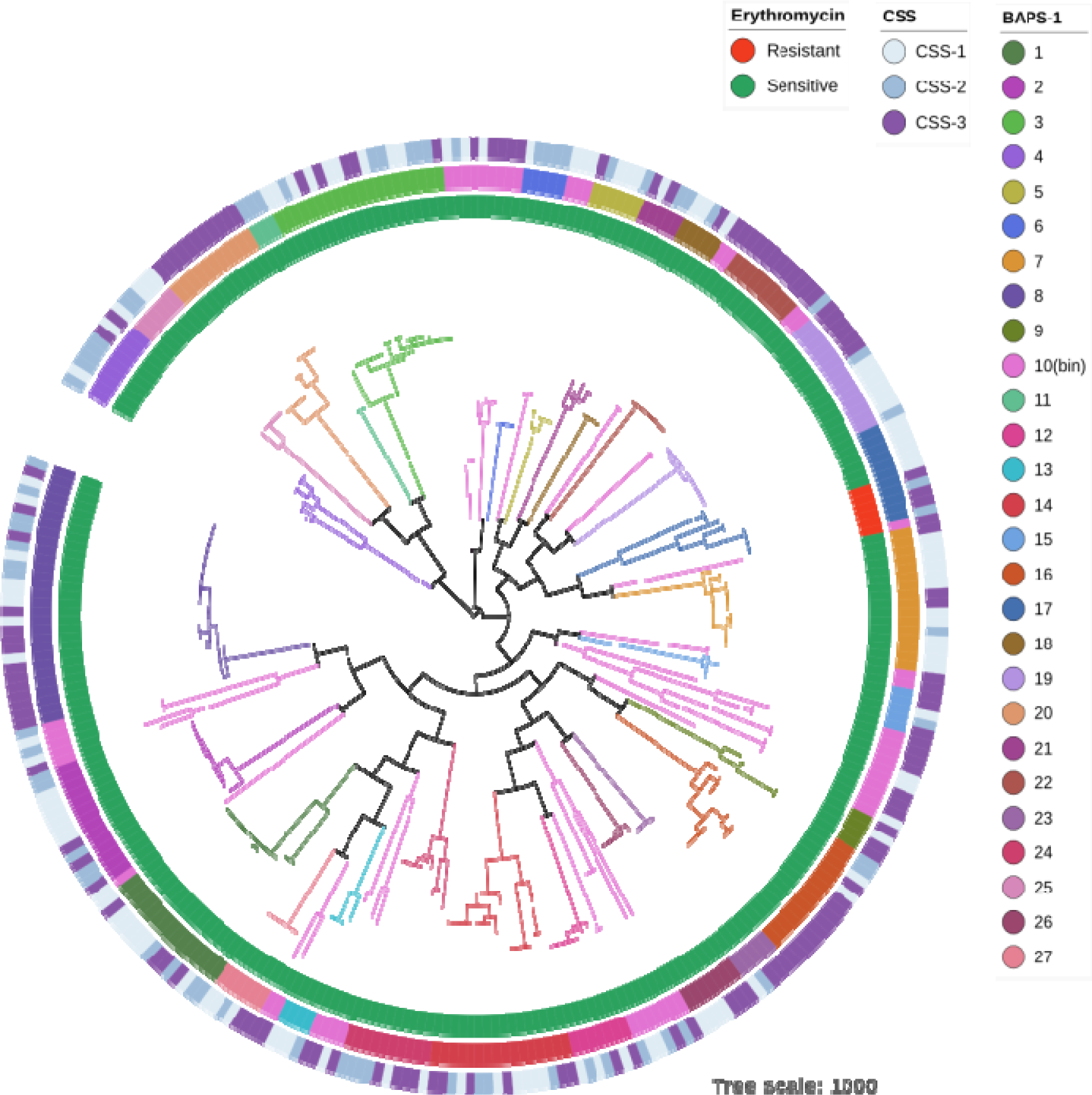
Phylogenetic tree using single nucleotide polymorphisms in the core genes of the 304 randomly sampled isolates. The innermost ring depicts each isolates susceptibility to erythromycin across the tree. The middle ring represents the BAPS clusters, of which BAPS10 (pink) is the bin cluster representing un-clustered isolates spread across the tree. The outer ring depicts the CSS in which the isolate was collected. The interactive tree is available at https://itol.embl.de/tree/193622058349121529412823.

At a finer resolution, sub-structure within the BAPS clusters was captured using a second round of BAPS clustering and identified 81 BAPS sub-clusters. None were observed to significantly change in proportion over the study period (Table 2).

**Table 2.** Proportion of isolates belonging to BAPS sub-clusters at each CSS.

| BAPS<br>sub-cluster | CSS-1 | CSS-2 | CSS-1 vs CSS-2 | CSS-3 | CSS-1 vs CSS-3 |
| --- | --- | --- | --- | --- | --- |
|  | Proportion (N=109) | Proportion (N=69) | p value | Proportion (N=126) | p value |
|  | % (95% CI) | % (95% CI) |  | % (95% CI) |  |
| 1 | 5.5 (2.05, 11.6) | 2.9 (0.35, 10.08) | 0.733 | 3.17 (0.87, 7.93) | 0.669 |
| 2 | 0.92 (0.02, 5.01) | 2.9 (0.35, 10.08) | NA | 0 (0, 2.89) | NA |
| 3 | 5.5 (2.05, 11.6) | 2.9 (0.35, 10.08) | 0.733 | 0 (0, 2.89) | 0.055 |
| 4 | 0 (0, 3.33) | 0 (0, 5.21) | NA | 0.79 (0.02, 4.34) | NA |
| 5 | 0 (0, 3.33) | 4.35 (0.91, 12.18) | NA | 1.59 (0.19, 5.62) | NA |
| 6 | 1.83 (0.22, 6.47) | 0 (0, 5.21) | NA | 0 (0, 2.89) | NA |
| 7 | 0.92 (0.02, 5.01) | 4.35 (0.91, 12.18) | NA | 2.38 (0.49, 6.8) | NA |
| 8 | 2.75 (0.57, 7.83) | 1.45 (0.04, 7.81) | NA | 1.59 (0.19, 5.62) | 0.748 |
| 9 | 1.83 (0.22, 6.47) | 2.9 (0.35, 10.08) | NA | 0 (0, 2.89) | NA |
| 10 | 0.92 (0.02, 5.01) | 0 (0, 5.21) | NA | 0 (0, 2.89) | NA |
| 11 | 0.92 (0.02, 5.01) | 4.35 (0.91, 12.18) | NA | 1.59 (0.19, 5.62) | NA |
| 12 | 0.92 (0.02, 5.01) | 2.9 (0.35, 10.08) | NA | 0 (0, 2.89) | NA |
| 13 | 0 (0, 3.33) | 1.45 (0.04, 7.81) | NA | 0.79 (0.02, 4.34) | NA |
| 14 | 2.75 (0.57, 7.83) | 1.45 (0.04, 7.81) | NA | 0 (0, 2.89) | NA |
| 15 | 0.92 (0.02, 5.01) | 1.45 (0.04, 7.81) | NA | 0 (0, 2.89) | NA |
| 16 | 0 (0, 3.33) | 4.35 (0.91, 12.18) | NA | 0 (0, 2.89) | NA |
| 17 | 0 (0, 3.33) | 0 (0, 5.21) | NA | 0 (0, 2.89) | NA |
| 18 | 5.5 (2.05, 11.6) | 1.45 (0.04, 7.81) | 0.733 | 0 (0, 2.89) | 0.055 |
| 19 | 3.67 (1.01, 9.13) | 0 (0, 5.21) | NA | 1.59 (0.19, 5.62) | 0.630 |
| 20 | 0 (0, 3.33) | 0 (0, 5.21) | NA | 0.79 (0.02, 4.34) | NA |
| 21 | 1.83 (0.22, 6.47) | 0 (0, 5.21) | NA | 0 (0, 2.89) | NA |
| 22 | 4.59 (1.51, 10.38) | 10.14 (4.18, 19.79) | 0.733 | 2.38 (0.49, 6.8) | 0.660 |
| 23 | 0 (0, 3.33) | 0 (0, 5.21) | NA | 4.76 (1.77, 10.08) | 0.095 |
| 24 | 2.75 (0.57, 7.83) | 0 (0, 5.21) | NA | 1.59 (0.19, 5.62) | 0.748 |
| 25 | 0 (0, 3.33) | 0 (0, 5.21) | NA | 2.38 (0.49, 6.8) | NA |
| 26 | 0 (0, 3.33) | 0 (0, 5.21) | NA | 1.59 (0.19, 5.62) | NA |
| 27 | 0 (0, 3.33) | 0 (0, 5.21) | NA | 0.79 (0.02, 4.34) | NA |
| 28 | 0.92 (0.02, 5.01) | 0 (0, 5.21) | NA | 0 (0, 2.89) | NA |
| 29 | 1.83 (0.22, 6.47) | 1.45 (0.04, 7.81) | NA | 0.79 (0.02, 4.34) | NA |
| 30 | 0 (0, 3.33) | 0 (0, 5.21) | NA | 1.59 (0.19, 5.62) | NA |
| 31 | 0.92 (0.02, 5.01) | 0 (0, 5.21) | NA | 3.17 (0.87, 7.93) | 0.630 |
| 32 | 3.67 (1.01, 9.13) | 1.45 (0.04, 7.81) | 0.754 | 4.76 (1.77, 10.08) | 0.800 |
| 33 | 2.75 (0.57, 7.83) | 0 (0, 5.21) | NA | 0 (0, 2.89) | NA |
| 34<sup>1</sup> | 4.59 (1.51, 10.38) | 7.25 (2.39, 16.11) | 0.733 | 10.32 (5.61, 17) | 0.279 |
| 35 | 0 (0, 3.33) | 1.45 (0.04, 7.81) | NA | 0 (0, 2.89) | NA |
| 36 | 0 (0, 3.33) | 1.45 (0.04, 7.81) | NA | 0 (0, 2.89) | NA |
| 37 | 0 (0, 3.33) | 1.45 (0.04, 7.81) | NA | 0 (0, 2.89) | NA |
| 38 | 0.92 (0.02, 5.01) | 0 (0, 5.21) | NA | 0.79 (0.02, 4.34) | NA |
| 39 | 1.83 (0.22, 6.47) | 2.9 (0.35, 10.08) | NA | 0.79 (0.02, 4.34) | NA |
| 40 | 2.75 (0.57, 7.83) | 0 (0, 5.21) | NA | 0 (0, 2.89) | NA |
| 41 | 0 (0, 3.33) | 1.45 (0.04, 7.81) | NA | 0 (0, 2.89) | NA |
| 42 | 2.75 (0.57, 7.83) | 4.35 (0.91, 12.18) | 0.754 | 3.17 (0.87, 7.93) | 1.000 |
| 43 | 0 (0, 3.33) | 0 (0, 5.21) | NA | 1.59 (0.19, 5.62) | NA |
| 44 | 2.75 (0.57, 7.83) | 1.45 (0.04, 7.81) | NA | 0 (0, 2.89) | NA |
| 46 | 0.92 (0.02, 5.01) | 1.45 (0.04, 7.81) | NA | 0.79 (0.02, 4.34) | NA |
| 47 | 0 (0, 3.33) | 0 (0, 5.21) | NA | 0.79 (0.02, 4.34) | NA |
| 48 | 0 (0, 3.33) | 0 (0, 5.21) | NA | 0.79 (0.02, 4.34) | NA |
| 49 | 0 (0, 3.33) | 0 (0, 5.21) | NA | 3.97 (1.3, 9.02) | 0.142 |
| 50 | 1.83 (0.22, 6.47) | 1.45 (0.04, 7.81) | NA | 0.79 (0.02, 4.34) | NA |
| 51 | 0 (0, 3.33) | 1.45 (0.04, 7.81) | NA | 0 (0, 2.89) | NA |
| 52 | 0 (0, 3.33) | 0 (0, 5.21) | NA | 3.17 (0.87, 7.93) | NA |
| 53 | 0 (0, 3.33) | 2.9 (0.35, 10.08) | NA | 2.38 (0.49, 6.8) | NA |
| 54 | 2.75 (0.57, 7.83) | 0 (0, 5.21) | NA | 0 (0, 2.89) | NA |
| 55 | 2.75 (0.57, 7.83) | 0 (0, 5.21) | NA | 0 (0, 2.89) | NA |
| 56 | 0.92 (0.02, 5.01) | 0 (0, 5.21) | NA | 0.79 (0.02, 4.34) | NA |
| 57 | 0.92 (0.02, 5.01) | 0 (0, 5.21) | NA | 0 (0, 2.89) | NA |
| 58 | 0.92 (0.02, 5.01) | 1.45 (0.04, 7.81) | NA | 0 (0, 2.89) | NA |
| 59 | 5.5 (2.05, 11.6) | 2.9 (0.35, 10.08) | 0.733 | 0 (0, 2.89) | 0.055 |
| 60 | 0 (0, 3.33) | 0 (0, 5.21) | NA | 4.76 (1.77, 10.08) | 0.095 |
| 61 | 0 (0, 3.33) | 0 (0, 5.21) | NA | 3.97 (1.3, 9.02) | 0.142 |
| 62 | 0 (0, 3.33) | 0 (0, 5.21) | NA | 1.59 (0.19, 5.62) | NA |
| 63 | 0 (0, 3.33) | 0 (0, 5.21) | NA | 3.17 (0.87, 7.93) | NA |
| 64 | 0.92 (0.02, 5.01) | 0 (0, 5.21) | NA | 0 (0, 2.89) | NA |
| 65 | 0 (0, 3.33) | 2.9 (0.35, 10.08) | NA | 0 (0, 2.89) | NA |
| 66 | 0 (0, 3.33) | 0 (0, 5.21) | NA | 0.79 (0.02, 4.34) | NA |
| 67 | 0 (0, 3.33) | 0 (0, 5.21) | NA | 0.79 (0.02, 4.34) | NA |
| 68 | 0 (0, 3.33) | 0 (0, 5.21) | NA | 5.56 (2.26, 11.11) | 0.072 |
| 69 | 0 (0, 3.33) | 0 (0, 5.21) | NA | 1.59 (0.19, 5.62) | NA |
| 70 | 0 (0, 3.33) | 0 (0, 5.21) | NA | 1.59 (0.19, 5.62) | NA |
| 71 | 0.92 (0.02, 5.01) | 1.45 (0.04, 7.81) | NA | 0 (0, 2.89) | NA |
| 72 | 0 (0, 3.33) | 0 (0, 5.21) | NA | 0.79 (0.02, 4.34) | NA |
| 73 | 0.92 (0.02, 5.01) | 5.8 (1.6, 14.18) | 0.733 | 1.59 (0.19, 5.62) | NA |
| 74 | 0.92 (0.02, 5.01) | 0 (0, 5.21) | NA | 0 (0, 2.89) | NA |
| 75 | 0 (0, 3.33) | 0 (0, 5.21) | NA | 1.59 (0.19, 5.62) | NA |
| 76 | 2.75 (0.57, 7.83) | 2.9 (0.35, 10.08) | 1.000 | 0 (0, 2.89) | NA |
| 77 | 0.92 (0.02, 5.01) | 0 (0, 5.21) | NA | 0 (0, 2.89) | NA |
| 78 | 3.67 (1.01, 9.13) | 0 (0, 5.21) | NA | 1.59 (0.19, 5.62) | 0.630 |
| 79 | 0 (0, 3.33) | 0 (0, 5.21) | NA | 0.79 (0.02, 4.34) | NA |
| 80 | 2.75 (0.57, 7.83) | 1.45 (0.04, 7.81) | NA | 0 (0, 2.89) | NA |
| 81 | 0 (0, 3.33) | 0 (0, 5.21) | NA | 1.59 (0.19, 5.62) | NA |

<sup>1</sup>bin cluster
NA =<5 isolates, not tested

### Serotype and sequence type distribution and diversity

Our analysis observed 36 serotypes and 81 STs. Fifteen STs were novel; 12 novel combinations of known alleles and 3 with 1 novel allele. None of the STs nor any of the serotypes were found to vary significantly in their proportion of the population during the study (Tables 3 and 4). At each CSS, serotype and ST diversity was high. No decrease in diversity was seen following the round of MDA, at either CSS-2 or CSS-3 (Table 5).

**Table 3.** Proportion of isolates belonging to a particular ST at each CSS.

| ST | CSS-1 | CSS-2 | CSS-1 vs CSS-2 | CSS-3 | CSS-1 vs CSS-3 |
| --- | --- | --- | --- | --- | --- |
|  | Proportion (N=109)<br>% (95% CI) | Proportion (N=69)<br>% (95% CI) | p value | Proportion (N=126)<br>% (95% CI) | p value |
| 63 | 0.92 (0.02, 5.01) | 0 (0, 5.21) | NA | 1.59 (0.19, 5.62) | NA |
| 177 | 0.92 (0.02, 5.01) | 0 (0, 5.21) | NA | 0 (0, 2.89) | NA |
| 180 | 0 (0, 3.33) | 0 (0, 5.21) | NA | 2.38 (0.49, 6.8) | NA |
| 260 | 0 (0, 3.33) | 0 (0, 5.21) | NA | 0.79 (0.02, 4.34) | NA |
| 361 | 2.75 (0.57, 7.83) | 1.45 (0.04, 7.81) | NA | 0 (0, 2.89) | NA |
| 447 | 0 (0, 3.33) | 0 (0, 5.21) | NA | 1.59 (0.19, 5.62) | NA |
| 458 | 2.75 (0.57, 7.83) | 0 (0, 5.21) | NA | 0 (0, 2.89) | NA |
| 802 | 0.92 (0.02, 5.01) | 1.45 (0.04, 7.81) | NA | 2.38 (0.49, 6.8) | NA |
| 847 | 5.5 (2.05, 11.6) | 7.25 (2.39, 16.11) | 1.000 | 2.38 (0.49, 6.8) | 0.653 |
| 910 | 5.5 (2.05, 11.6) | 2.9 (0.35, 10.08) | 0.974 | 3.17 (0.87, 7.93) | 0.728 |
| 913 | 0.92 (0.02, 5.01) | 5.8 (1.6, 14.18) | 0.375 | 1.59 (0.19, 5.62) | NA |
| 915 | 2.75 (0.57, 7.83) | 1.45 (0.04, 7.81) | NA | 1.59 (0.19, 5.62) | 0.776 |
| 925 | 0.92 (0.02, 5.01) | 1.45 (0.04, 7.81) | NA | 0 (0, 2.89) | NA |
| 989 | 0 (0, 3.33) | 1.45 (0.04, 7.81) | NA | 0 (0, 2.89) | NA |
| 1469 | 2.75 (0.57, 7.83) | 2.9 (0.35, 10.08) | 1.000 | 0 (0, 2.89) | NA |
| 1735 | 0 (0, 3.33) | 0 (0, 5.21) | NA | 0.79 (0.02, 4.34) | NA |
| 1736 | 0.92 (0.02, 5.01) | 0 (0, 5.21) | NA | 0 (0, 2.89) | NA |
| 1737 | 0 (0, 3.33) | 0 (0, 5.21) | NA | 1.59 (0.19, 5.62) | NA |
| 1778 | 3.67 (1.01, 9.13) | 0 (0, 5.21) | NA | 1.59 (0.19, 5.62) | 0.653 |
| 1794 | 0 (0, 3.33) | 2.9 (0.35, 10.08) | NA | 2.38 (0.49, 6.8) | NA |
| 2169 | 0 (0, 3.33) | 0 (0, 5.21) | NA | 0.79 (0.02, 4.34) | NA |
| 2174 | 0 (0, 3.33) | 0 (0, 5.21) | NA | 1.59 (0.19, 5.62) | NA |
| 2178 | 0.92 (0.02, 5.01) | 1.45 (0.04, 7.81) | NA | 0.79 (0.02, 4.34) | NA |
| 2831 | 0 (0, 3.33) | 0 (0, 5.21) | NA | 7.14 (3.32, 13.13) | 0.056 |
| 3310 | 0 (0, 3.33) | 0 (0, 5.21) | NA | 0.79 (0.02, 4.34) | NA |
| 3329 | 1.83 (0.22, 6.47) | 2.9 (0.35, 10.08) | NA | 4.76 (1.77, 10.08) | 0.653 |
| 3404 | 0.92 (0.02, 5.01) | 2.9 (0.35, 10.08) | NA | 0 (0, 2.89) | NA |
| 3406 | 0 (0, 3.33) | 0 (0, 5.21) | NA | 1.59 (0.19, 5.62) | NA |
| 3407 | 8.26 (3.85, 15.1) | 8.7 (3.26, 17.97) | 1.000 | 2.38 (0.49, 6.8) | 0.248 |
| 4033 | 7.34 (3.22, 13.95) | 8.7 (3.26, 17.97) | 1.000 | 11.11 (6.21, 17.94) | 0.653 |
| 4832 | 0 (0, 3.33) | 0 (0, 5.21) | NA | 0.79 (0.02, 4.34) | NA |
| 4989 | 0 (0, 3.33) | 1.45 (0.04, 7.81) | NA | 0.79 (0.02, 4.34) | NA |
| 5103 | 0.92 (0.02, 5.01) | 2.9 (0.35, 10.08) | NA | 0 (0, 2.89) | NA |
| 5349 | 0.92 (0.02, 5.01) | 1.45 (0.04, 7.81) | NA | 2.38 (0.49, 6.8) | NA |
| 5505 | 2.75 (0.57, 7.83) | 1.45 (0.04, 7.81) | NA | 0.79 (0.02, 4.34) | NA |
| 5511 | 0 (0, 3.33) | 0 (0, 5.21) | NA | 0.79 (0.02, 4.34) | NA |
| 5515 | 0.92 (0.02, 5.01) | 0 (0, 5.21) | NA | 0 (0, 2.89) | NA |
| 5516 | 0 (0, 3.33) | 1.45 (0.04, 7.81) | NA | 0 (0, 2.89) | NA |
| 5521 | 5.5 (2.05, 11.6) | 2.9 (0.35, 10.08) | 0.974 | 4.76 (1.77, 10.08) | 1.000 |
| 5702 | 2.75 (0.57, 7.83) | 2.9 (0.35, 10.08) | 1.000 | 1.59 (0.19, 5.62) | 0.776 |
| 5706 | 0.92 (0.02, 5.01) | 0 (0, 5.21) | NA | 0 (0, 2.89) | NA |
| 5719 | 3.67 (1.01, 9.13) | 0 (0, 5.21) | NA | 1.59 (0.19, 5.62) | 0.653 |
| 5722 | 0 (0, 3.33) | 1.45 (0.04, 7.81) | NA | 0 (0, 2.89) | NA |
| 5726 | 1.83 (0.22, 6.47) | 0 (0, 5.21) | NA | 0 (0, 2.89) | NA |
| 5734 | 0.92 (0.02, 5.01) | 4.35 (0.91, 12.18) | NA | 1.59 (0.19, 5.62) | NA |
| 5823 | 0 (0, 3.33) | 0 (0, 5.21) | NA | 0.79 (0.02, 4.34) | NA |
| 5902 | 2.75 (0.57, 7.83) | 0 (0, 5.21) | NA | 0 (0, 2.89) | NA |
| 7336 | 0 (0, 3.33) | 1.45 (0.04, 7.81) | NA | 0.79 (0.02, 4.34) | NA |
| 8949 | 0 (0, 3.33) | 0 (0, 5.21) | NA | 5.56 (2.26, 11.11) | 0.074 |
| 10542 | 0 (0, 3.33) | 0 (0, 5.21) | NA | 1.59 (0.19, 5.62) | NA |
| 10593 | 2.75 (0.57, 7.83) | 0 (0, 5.21) | NA | 0 (0, 2.89) | NA |
| 10607 | 0 (0, 3.33) | 2.9 (0.35, 10.08) | NA | 0 (0, 2.89) | NA |
| 10610 | 0 (0, 3.33) | 0 (0, 5.21) | NA | 1.59 (0.19, 5.62) | NA |
| 10679 | 5.5 (2.05, 11.6) | 1.45 (0.04, 7.81) | 0.835 | 0 (0, 2.89) | 0.065 |
| 10777 | 0 (0, 3.33) | 0 (0, 5.21) | NA | 3.17 (0.87, 7.93) | NA |
| 10963 | 1.83 (0.22, 6.47) | 0 (0, 5.21) | NA | 0 (0, 2.89) | NA |
| 11707 | 0 (0, 3.33) | 0 (0, 5.21) | NA | 0.79 (0.02, 4.34) | NA |
| 11710 | 0 (0, 3.33) | 0 (0, 5.21) | NA | 0.79 (0.02, 4.34) | NA |
| 11720 | 0.92 (0.02, 5.01) | 5.8 (1.6, 14.18) | 0.375 | 0 (0, 2.89) | NA |
| 11756 | 0.92 (0.02, 5.01) | 0 (0, 5.21) | NA | 0 (0, 2.89) | NA |
| 11758 | 0 (0, 3.33) | 0 (0, 5.21) | NA | 0.79 (0.02, 4.34) | NA |
| 12287 | 2.75 (0.57, 7.83) | 1.45 (0.04, 7.81) | NA | 0 (0, 2.89) | NA |
| 12288 | 0 (0, 3.33) | 4.35 (0.91, 12.18) | NA | 0 (0, 2.89) | NA |
| 12314 | 0 (0, 3.33) | 0 (0, 5.21) | NA | 2.38 (0.49, 6.8) | NA |
| 12692 | 0.92 (0.02, 5.01) | 0 (0, 5.21) | NA | 0.79 (0.02, 4.34) | NA |
| 13845 | 0 (0, 3.33) | 0 (0, 5.21) | NA | 0.79 (0.02, 4.34) | NA |
| 13846 | 0.92 (0.02, 5.01) | 0 (0, 5.21) | NA | 0 (0, 2.89) | NA |
| 13847 | 0.92 (0.02, 5.01) | 1.45 (0.04, 7.81) | NA | 0 (0, 2.89) | NA |
| 13848 | 0 (0, 3.33) | 0 (0, 5.21) | NA | 1.59 (0.19, 5.62) | NA |
| 13849 | 0.92 (0.02, 5.01) | 0 (0, 5.21) | NA | 0 (0, 2.89) | NA |
| 13850 | 0.92 (0.02, 5.01) | 0 (0, 5.21) | NA | 0 (0, 2.89) | NA |
| 13850 | 0.92 (0.02, 5.01) | 0 (0, 5.21) | NA | 0 (0, 2.89) | NA |
| 13852 | 0 (0, 3.33) | 0 (0, 5.21) | NA | 1.59 (0.19, 5.62) | NA |
| 13853 | 1.83 (0.22, 6.47) | 1.45 (0.04, 7.81) | NA | 2.38 (0.49, 6.8) | 1.000 |
| 13854 | 0 (0, 3.33) | 0 (0, 5.21) | NA | 0.79 (0.02, 4.34) | NA |
| 13855 | 0.92 (0.02, 5.01) | 2.9 (0.35, 10.08) | NA | 0 (0, 2.89) | NA |
| 13856 | 1.83 (0.22, 6.47) | 0 (0, 5.21) | NA | 0 (0, 2.89) | NA |
| 13857 | 0.92 (0.02, 5.01) | 1.45 (0.04, 7.81) | NA | 0.79 (0.02, 4.34) | NA |
| 13858 | 0 (0, 3.33) | 0 (0, 5.21) | NA | 3.17 (0.87, 7.93) | NA |
| 13859 | 0 (0, 3.33) | 1.45 (0.04, 7.81) | NA | 0 (0, 2.89) | NA |

NA =<5 isolates, not tested

**Table 4.** Proportion of isolates belonging to a particular serotype at each CSS.

| Serotype | CSS-1 | CSS-2 | CSS-1 vs CSS-2 | CSS-3 | CSS-1 vs CSS-3 |
| --- | --- | --- | --- | --- | --- |
|  | Proportion (N=109) | Proportion (N=69) | p value | Proportion (N=126) | p value |
|  | % (95% CI) | % (95% CI) |  | % (95% CI) |  |
| 3 | 4.59 (1.51, 10.38) | 0 (0, 5.21) | 0.644 | 3.17 (0.87, 7.93) | 1.000 |
| 4 | 3.67 (1.01, 9.13) | 2.9 (0.35, 10.08) | 1.000 | 0 (0, 2.89) | NA |
| 5 | 0.92 (0.02, 5.01) | 2.9 (0.35, 10.08) | NA | 0 (0, 2.89) | NA |
| 6A | 5.5 (2.05, 11.6) | 11.59 (5.14, 21.57) | 0.644 | 4.76 (1.77, 10.08) | 1.000 |
| 6B | 0.92 (0.02, 5.01) | 2.9 (0.35, 10.08) | NA | 0 (0, 2.89) | NA |
| 7F | 0.92 (0.02, 5.01) | 1.45 (0.04, 7.81) | NA | 0.79 (0.02, 4.34) | NA |
| 8 | 0 (0, 3.33) | 0 (0, 5.21) | NA | 1.59 (0.19, 5.62) | NA |
| 9L | 0 (0, 3.33) | 0 (0, 5.21) | NA | 0.79 (0.02, 4.34) | NA |
| 9V | 3.67 (1.01, 9.13) | 0 (0, 5.21) | NA | 2.38 (0.49, 6.8) | 1.000 |
| 10A | 5.5 (2.05, 11.6) | 2.9 (0.35, 10.08) | 0.974 | 4.76 (1.77, 10.08) | 1.000 |
| 10B | 2.75 (0.57, 7.83) | 1.45 (0.04, 7.81) | NA | 0.79 (0.02, 4.34) | NA |
| 11A | 2.75 (0.57, 7.83) | 2.9 (0.35, 10.08) | 1.000 | 0 (0, 2.89) | NA |
| 12F | 0 (0, 3.33) | 1.45 (0.04, 7.81) | NA | 0 (0, 2.89) | NA |
| 13 | 0.92 (0.02, 5.01) | 5.8 (1.6, 14.18) | 0.5997 | 0.79 (0.02, 4.34) | NA |
| 14 | 3.67 (1.01, 9.13) | 1.45 (0.04, 7.81) | 1.000 | 3.97 (1.3, 9.02) | 1.000 |
| 15B/15C | 12.84 (7.2, 20.61) | 11.59 (5.14, 21.57) | 1.000 | 12.7 (7.44, 19.8) | 1.000 |
| 16F | 9.17 (4.49, 16.23) | 13.04 (6.14, 23.32) | 0.974 | 12.7 (7.44, 19.8) | 1.000 |
| 17F | 2.75 (0.57, 7.83) | 0 (0, 5.21) | NA | 0.79 (0.02, 4.34) | NA |
| 18C | 0.92 (0.02, 5.01) | 0 (0, 5.21) | NA | 0 (0, 2.89) | NA |
| 19A | 5.5 (2.05, 11.6) | 7.25 (2.39, 16.11) | 1.000 | 4.76 (1.77, 10.08) | 1.000 |
| 19B | 2.75 (0.57, 7.83) | 4.35 (0.91, 12.18) | 1.000 | 3.17 (0.87, 7.93) | 1.000 |
| 19F | 5.5 (2.05, 11.6) | 5.8 (1.6, 14.18) | 1.000 | 3.17 (0.87, 7.93) | 1.000 |
| 20 | 1.83 (0.22, 6.47) | 2.9 (0.35, 10.08) | NA | 2.38 (0.49, 6.8) | 1.000 |
| 21 | 1.83 (0.22, 6.47) | 4.35 (0.91, 12.18) | 0.974 | 1.59 (0.19, 5.62) | NA |
| 23A | 2.75 (0.57, 7.83) | 2.9 (0.35, 10.08) | 1.000 | 0.79 (0.02, 4.34) | NA |
| 23B | 0.92 (0.02, 5.01) | 0 (0, 5.21) | NA | 0.79 (0.02, 4.34) | NA |
| 23F | 1.83 (0.22, 6.47) | 4.35 (0.91, 12.18) | 0.974 | 3.17 (0.87, 7.93) | 1.000 |
| 24 | 0 (0, 3.33) | 0 (0, 5.21) | NA | 2.38 (0.49, 6.8) | NA |
| 28F | 0 (0, 3.33) | 0 (0, 5.21) | NA | 0.79 (0.02, 4.34) | NA |
| 33D | 0.92 (0.02, 5.01) | 0 (0, 5.21) | NA | 3.17 (0.87, 7.93) | 1.000 |
| 34 | 11.01 (5.82, 18.44) | 1.45 (0.04, 7.81) | 0.280 | 3.97 (1.3, 9.02) | 0.239 |
| 37 | 1.83 (0.22, 6.47) | 2.9 (0.35, 10.08) | NA | 7.94 (3.87, 14.11) | 0.239 |
| 35B | 0.92 (0.02, 5.01) | 1.45 (0.04, 7.81) | NA | 2.38 (0.49, 6.8) | NA |
| 35F | 0 (0, 3.33) | 0 (0, 5.21) | NA | 2.38 (0.49, 6.8) | NA |
| 38 | 0.92 (0.02, 5.01) | 0 (0, 5.21) | NA | 0 (0, 2.89) | NA |
| 45 | 0 (0, 3.33) | 0 (0, 5.21) | NA | 7.14 (3.32, 13.13) | 0.064 |

NA =<5 isolates, not tested

**Table 5.** Serotype and ST diversity at each CSS.

|  | CSS-1 | CSS-2 | CSS-3 |
| --- | --- | --- | --- |
| Serotype<sup>1</sup> | 0.94 | 0.93 | 0.94 |
| ST<sup>1</sup> | 0.96 | 0.96 | 0.96 |

<sup>1</sup>Diversity was assessed using Simpsons Diversity index (Dominance Index Approximation), which represents the probability that two randomly selected strains are the same, where 1 means they are different and 0 indicates they are the same.

### Antimicrobial resistance

Five isolates, all serotype 20, ST1794 strains of BAPS17, carried the macrolide efflux genes *mefA* and msrD, which confer macrolide resistance^25^ (Figure 1). These were found together in a Tn2010 element, which also carried tetM, conferring tetracyline resistance

[23]. Each of the isolates also had mutations in *FolA* and *FolP*, which confer resistance to co-trimoxazole. These multi-drug resistant strains were isolated at both CSS-2 (2 isolates) and CSS-3 (3 isolates). The proportion of isolates predicted to be erythromycin-, tetracycline- and co-trimoxazole-resistant within BAPS17 therefore increased significantly between CSS-1 (0 of 6 BAPS17 isolates) and CSS-2 (3 of 3 BAPS17 isolates) / CCS-3 (2 of 2 BAPS17 isolates) (p<0.05). Only one additional isolate, belonging to serotype 13, carried the macrolide efflux gene *mefE* [22].

## DISCUSSION

MDA with azithromycin for trachoma elimination in The Gambia was associated with a marked but short-term decrease in the prevalence of nasopharyngeal S. *pneumoniae* carriage ^9^. In an effort to evaluate whether communities were repopulated with the same strains following treatment or whether MDA gave rise to previously suppressed clones, we compared S. *pneumoniae* isolates obtained from communities sampled before and after a third round of MDA. Our results show that in communities where the number of circulating lineages was high and the prevalence of macrolide resistance was low, little change is seen in population structure following a round of MDA.

The number of BAPS clusters identified in our dataset was higher than other published carriage datasets using this approach, which likely reflects the broader pneumococcal diversity in The Gambia with small numbers of many different lineages circulating^13,14^. No changes in BAPS clusters were identified at CSS-2 suggesting any change that MDA had on the population structure was short-lived, or that these changes were below our detection limit. Greater variation was seen between CSS-1 and CSS-3, six months following MDA. These changes could be a result of natural temporal fluctuations^26^ or could be a reflection of seasonal change ^27^; CSS-1 and CCS-2 were conducted in the wet season while CSS-3 was carried out in the dry season. Only two BAPS clusters, BAPS20 and 22, showed an increase between CSS-1 and CSS-3. However these changes were not associated with macrolide resistance and are difficult to attribute to the MDA.

Macrolide resistance conferred by *ermB* in this sample-set was confined to BAPS17, this lineage did not increase significantly following treatment. However, the proportion of macrolide resistance isolates within BAPS17 was higher at both CSS-2 and CSS-3 indicating an increase after MDA. We are unable rule out the presence of macrolide resistant BAPS17 strains at CSS-1 due to the relatively small sample size in the background of high ST diversity. It may be that sensitive ST1794 strains of BAPS17 initially supressed resistant strains and that these expanded after treatment. Alternatively, strains may have acquired resistance after MDA through a recombination event with other pneumococci. ST1794 was only observed once in the published international dataset ^28^ despite inclusion of 1249 carriage isolates from The Gambia. All isolates of BAPS17 belong to the Global Pneumococcal Sequence Cluster GPSC61. Within GPSC61 *ermB* and/or *mef* were not detected in the single ST1794 representative or other STs, therefore GPSC61 is not a lineage that was associated with macrolide resistance. Although the numbers are small, all isolates that carried macrolide resistance genes concurrently carried tetracycline- and co-trimoxazole macrolide resistance genes in mobile genetic elements. If MDA selects for strains carrying mobile genetic elements which do often encode more than one antibiotic resistance gene, there is potential for increases in multi-drug resistance ^29^. Macrolide resistance was at undetectable levels in this study before the intervention, in other settings with established macrolide resistance in pneumococci, the potential for rises in macrolide resistant pneumococci after MDA with azithromycin may be greater. Further studies with larger sample sizes would be needed to adequately address this concern and would be important if azithromycin MDA were to be considered as an intervention to reduce child mortality^5^.

### Limitations

High serotype diversity was seen both before and after MDA suggesting that, while the exact serotypes found at a given time-point may vary, the number of different serotypes found remains unchanged. However, our study was limited in its power to detect changes in serotype and ST distribution given the small sample size relative to the high pneumococcal diversity observed. And whilst multiple colonies with varying morphologies were archived from each participant during the initial pneumococcal carriage study, sequencing revealed these predominantly represented the same serotype and ST. After duplicated isolates were excluded from statistical analyses (as they were not independent isolates), the resulting sample size prevented us from detecting small changes (two-or three-fold increases in BAPS frequency) following MDA.

Whole genome sequencing was carried out on a random selection of isolates generated from our previous pneumococcal carriage surveys however, only 75% of selected isolates were eventually sequenced. This was due to a number of isolates that either could not be revived from glycerol stocks, or did not grow to sufficient density in Todd Hewitt enrichment broth to enable extraction of the required quantity of DNA. This may have resulted in an underrepresentation of serotypes or STs that are slow-growing or otherwise less robust.

Due to the study design, we did not have isolates obtained before any MDA took place in our study villages. We therefore cannot rule out that the three-year MDA programme, as a whole, did not affect S. *pneumoniae* population structure, although it clearly did not lead to a substantial increase in the proportion of macrolide resistant strains as only 6/304 (2%) independent isolates studied here carried macrolide resistance genes. We screened for known genetic determinants of macrolide resistance therefore novel mechanisms or efflux related tolerance will not have been detected. However, the detection of genetic determinants *mef* and/or *ermB* has been reported to have a sensitivity of 97% and specificity of 99% for phenotypic resistance^28^.

### Conclusions

Few changes in S. *pneumoniae* population structure were observed between CSS-1 and CSS-3, before and six months following the third round of MDA however the study was limited by lack of power to detect small fold changes in BAPS clustering. While a small number of isolates possessed genes encoding macrolide resistance, these were found on a mobile genetic element carrying another antimicrobial resistance element. Further studies with larger sample sizes would be needed to determine whether azithromycin MDA selects for multi-drug resistance strains.

## Data Availability

Data supporting the findings is contained within the manuscript. An interactive phylogenetic tree is available at itol
Metadata including ENA accession numbers and phylogeny are deposited in Microreact.

https://itol.embl.de/tree/193622058349121529412823.

https://microreact.org/project/MDAandpneumo

## FUNDING

This work was supported by the Bill and Melinda Gates Foundation [grant number 48027], by the Wellcome Trust [grant number WT093368MA] and by Wellcome Trust core funding to the Wellcome Trust Sanger Institute [grant number 206194]. The funders played no role in the design, collection, analysis and interpretation of the data, in the writing of the manuscript and in the decision to send the manuscript for publication.

## ACKNOWLEDGEMENTS

We are grateful to the community leaders and villagers for their participation in the study.

All authors declare no conflicting interests.

## SUPPLEMENTARY DATA

All supplementary data including a phylogenetic tree of all 514 isolates sequenced and ENA accession numbers are deposited in Microreact https://microreact.org/project/MDAandpneumo.

## Notes

### Competing Interest Statement

The authors have declared no competing interest.

### Clinical Trial

This study was ancillary to the Partnership for the Rapid Elimination of Trachoma, ClinicalTrials.gov NCT00792922, registration date November 17, 2008.

